# Evaluation of COVID-19 vaccination strategies with a delayed second dose

**DOI:** 10.1101/2021.01.27.21250619

**Authors:** Seyed M. Moghadas, Thomas N. Vilches, Kevin Zhang, Shokoofeh Nourbakhsh, Pratha Sah, Meagan C. Fitzpatrick, Alison P. Galvani

## Abstract

Two of the COVID-19 vaccines currently approved in the United States require two doses, administered three to four weeks apart. Constraints in vaccine supply and distribution capacity, together with a deadly wave of COVID-19 from November 2020 to January 2021 and the emergence of highly contagious SARS-CoV-2 variants, sparked a policy debate on whether to vaccinate more individuals with the first dose of available vaccines and delay the second dose, or to continue with the recommended two-dose series as tested in clinical trials. We developed an agent-based model of COVID-19 transmission to compare the impact of these two vaccination strategies, while varying the temporal waning of vaccine efficacy following the first dose and the level of pre-existing immunity in the population. Our results show that for Moderna vaccines, a delay of at least 9 weeks could maximize vaccination program effectiveness and avert at least an additional 17.3 (95% CrI: 7.8 − 29.7) infections, 0.71 (95% CrI: 0.52 - 0.97) hospitalizations, and 0.34 (95% CrI: 0.25 - 0.44) deaths per 10,000 population compared to the recommended 4-week interval between the two doses. Pfizer-BioNTech vaccines also averted an additional 0.61 (95% CrI: 0.37 - 0.89) hospitalizations and 0.31 (95% CrI: 0.23 - 0.45) deaths per 10,000 population in a 9-week delayed second dose strategy compared to the 3-week recommended schedule between doses. However, there was no clear advantage of delaying the second dose with Pfizer-BioNTech vaccines in reducing infections, unless the efficacy of the first dose did not wane over time. Our findings underscore the importance of quantifying the characteristics and durability of vaccine-induced protection after the first dose in order to determine the optimal time interval between the two doses.

## Introduction

The spread of coronavirus disease 2019 (COVID-19) has ravaged global health and suppressed economic activity despite the range of mitigation measures implemented by countries worldwide [1]. A number of vaccines, including those developed by Pfizer-BioNTech, Moderna, and Oxford-AstraZeneca, have received emergency use authorization from regulatory bodies in different countries [2]. Clinical trials and evaluations of mass vaccination campaigns have demonstrated that these vaccines can provide high levels of protection against symptomatic and severe disease with two doses administered three to four weeks apart [3–6]. In contrast to the remarkable speed of development, vaccine delivery has proven to be challenging due to supply shortages and limited distribution capacity in several countries [7,8].

The emergence of novel, more contagious SARS-CoV-2 variants in several countries [9–13], and the potential for their widespread transmission have led to a public health conundrum regarding whether to vaccinate more individuals with the first dose of available vaccines and delay the second dose, or to prioritize completion of the two-dose series based on tested schedules in clinical trials [14–17]. Broader population-level protection against COVID-19 in a delayed second dose (DSD) strategy, even with lower individual-level efficacy from the first dose in the short term, may improve the impact of vaccination compared to the recommended two-dose strategy that provides more complete protection to a smaller subset of the population [14,17].

However, the conditions under which this improvement is achievable remain unexamined [18], such as the durability of first-dose efficacy and protection against infection [19–21].

Here, we employed an agent-based model of COVID-19 transmission and vaccination to compare the epidemiological impact of tested and DSD vaccination schedules, considering a range of pre-existing immunity accrued since the emergence of COVID-19. We determined the optimal timing for administering the second dose based on vaccine efficacy estimated in clinical trials and population-level studies following first and second doses [3,4,6,22–24]. For Moderna’s two-dose vaccine, we show that a DSD strategy would outperform the recommended interval between doses in terms of reducing the number of hospitalizations and deaths. The maximum benefits would be achieved with a delay of at least 9 weeks from the recommended schedule for administering the second dose. A DSD strategy with Pfizer-BioNTech vaccines is comparatively inferior to Moderna vaccines, and the delay to achieve maximum benefits depends on the durability of the first-dose efficacy.

## Methods

### Model structure

We extended our previous model of agent-based COVID-19 transmission to include vaccination [25]. The model encapsulates the natural history of COVID-19 with classes of individuals including: susceptible; vaccinated; latently infected (not yet infectious); asymptomatic (and infectious); pre-symptomatic (and infectious); symptomatic with either mild or severe illness; recovered; and dead. Model population was stratified into six age groups of 0-4, 5-19, 20-49, 50-64, 65-79, and 80+ years based on United States (US) census data [26]. We sampled daily contacts within and between age groups from a negative-binomial distribution parameterized using an empirically-determined contact network (Appendix, Table A1) [27].

### Disease dynamics

In our agent-based model, the risk of infection for people susceptible to COVID-19 depended on contact with infectious individuals that could be in asymptomatic, pre-symptomatic, or symptomatic stages of the disease. Using recent estimates, we parameterized the infectivity of asymptomatic, mild symptomatic, and severe symptomatic individuals to be 26%, 44%, and 89% relative to the pre-symptomatic stage [28–30]. For each infected individual, the incubation period was sampled from a Gamma distribution with a mean of 5.2 days [31]. A proportion of infected individuals developed symptomatic disease following a pre-symptomatic stage. The duration of the pre-symptomatic stage and infectious period following symptom onset was sampled from a Gamma distribution with a mean of 2.3 days and 3.2 days, respectively [29,32,33]. Those who did not develop symptomatic disease remained asymptomatic until recovery, with an infectious period that was sampled from a Gamma distribution with a mean of 5 days [33,34]. We assumed that recovered individuals are immune against reinfection for the remainder of simulation timelines. Model parameters are summarized in Appendix, Table A2.

### Infection outcomes

A proportion of severe symptomatic cases were hospitalized within 2-5 days of symptom onset [25,35], and thereafter did not contribute to the spread of infection. We assumed that all mild symptomatic cases and severely ill individuals self-isolated within 24 hours of symptom onset. The daily number of contacts during self-isolation was reduced by an average of 74%, based on a matrix derived from a representative sample population during COVID-19 lockdown [36]. Non-ICU and ICU admissions were parameterized based on age-stratified data for COVID-19 hospitalizations, and the presence of comorbidities [37,38]. The lengths of non-ICU and ICU stays were sampled from Gamma distributions with means of 12.4 and 14.4 days, respectively [39,40].

### Vaccination

We implemented a two-dose vaccination campaign, and simulated a rollout strategy with a daily rate of 30 vaccine doses per 10,000 population, corresponding to 1 million vaccine doses per day for the entire US population. This rate corresponds to the goal of administering ∼100 million vaccine doses in the first 100 days, as outlined by the Biden administration [33]. As an additional scenario, we extended our analysis to a daily vaccination rate of 45 doses per 10,000 population, corresponding to 1.5 million daily doses in the US. In all scenarios, prioritization was sequentially set to: (i) healthcare workers (5% of the total population) [41], adults with comorbidities, and those aged 65 and older; and (ii) other individuals aged 18-64 [42]. We assumed that the maximum achievable coverage was 95% among healthcare workers and those aged 65 and older. This maximum coverage was set to 70% among other age groups, with an age-dependent distribution (Appendix, Table A3).

For Moderna vaccines, the interval between the first and second doses in the recommended schedule (i.e., tested in clinical trials) was 28 days [3]. This interval was 21 days for Pfizer-BioNTech vaccines [4]. Vaccine coverage of the entire population with two doses under the recommended schedules reached 51% and 76% for Moderna, and 52% and 77% for Pfizer-BioNTech with vaccination rates of 30 and 45 doses per day, respectively, within one year.

We performed a review of published studies and the US Food and Drug Administration (FDA) briefing documents on the efficacy of Moderna and Pfizer-BioNTech vaccines in preventing infection, symptomatic disease, and severe disease [3,4,6,22–24]. We extracted reported estimates for vaccine efficacies and associated timelines, as presented in Figure 1. These estimates indicate no statistically significant difference between the protection of vaccinated and unvaccinated cohorts for the first 10-14 days following the first dose of vaccines.

**Figure 1.**
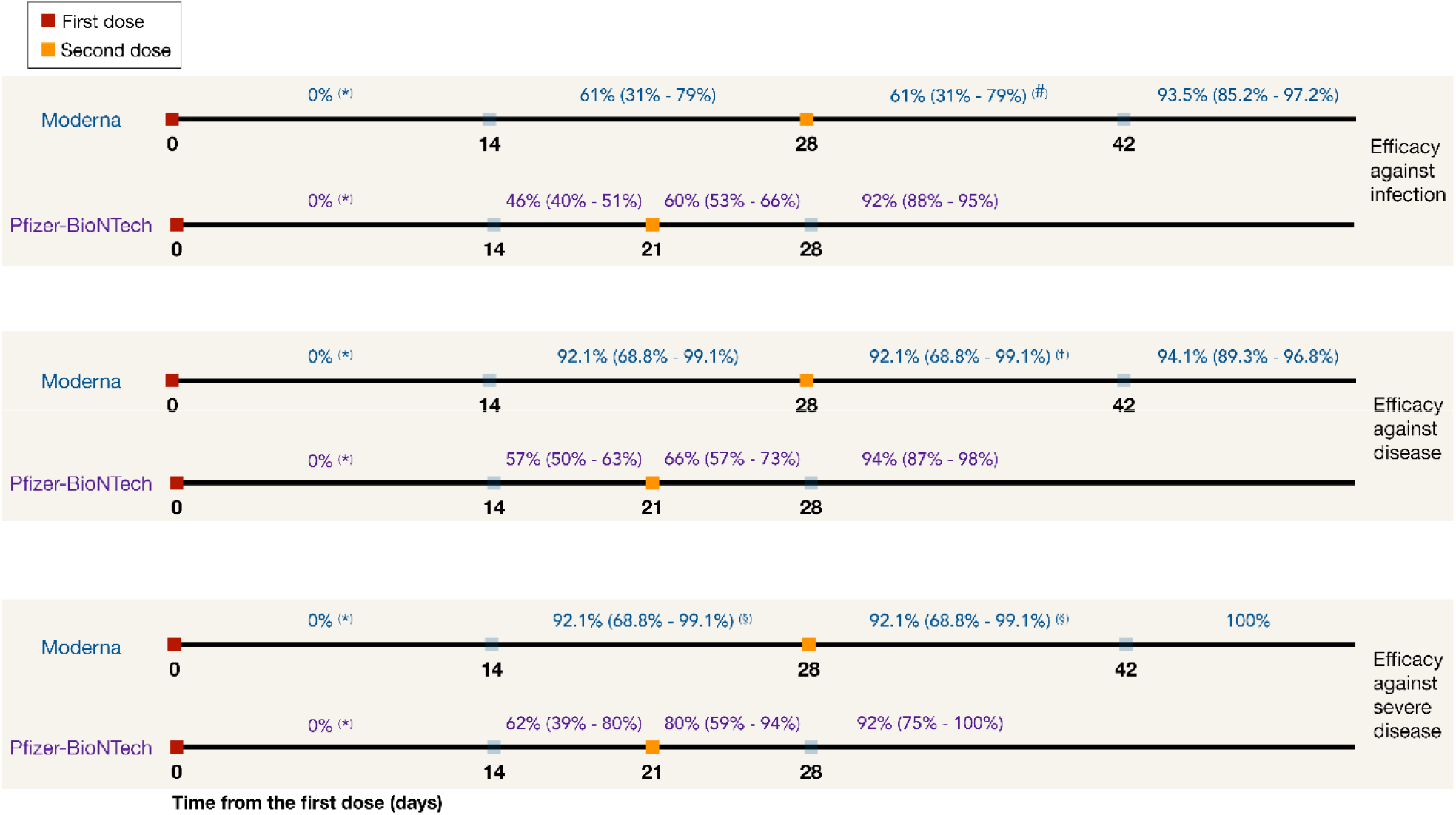
Efficacy of Moderna and Pfizer-BioNTech vaccines against all infection, symptomatic disease and severe disease, derived from published studies and US FDA briefing documents [3,4,6,22–24]. (*) During the first 14 days following the first dose of vaccines, there was no statistically significant difference between the protection in the vaccinated and unvaccinated cohorts. (#) Conservatively assumed to be the same as efficacy against infection during the preceding 14 days (prior to the second dose). (†) Conservatively assumed to be the same as efficacy against severe disease during the preceding 14 days (prior to the second dose). (§) Assumed to be the same as efficacy against symptomatic COVID-19.

In order to evaluate the impact of vaccination with DSD relative to the tested schedules in clinical trials, in the base-case scenario, we assumed that the efficacy of the first dose for both Moderna and Pfizer-BioNTech vaccines would be maintained for up to 18 weeks without a second dose. As a sensitivity analysis, we considered a waning rate of 5% per week for first-dose vaccine efficacy starting from week 7 after the first dose. We assumed that the full two-dose efficacy was achieved regardless of delay in the administration of the second dose from the recommended schedule [43]. We simulated the model with the mean, lower bound, and upper bound of the 95% CIs for vaccine efficacy of the first and second doses against infection, symptomatic disease, and severe disease.

### Model scenarios

We considered a range of 10%-30% pre-existing immunity (i.e., seropositivity prior to vaccination) in the population, with 20% for the base-case scenario [44,45]. To parameterize the model at a given level of pre-existing immunity, we ran simulations in the absence of vaccination, and derived the infection rates in different age groups once the overall attack rate reached the pre-specified level. The corresponding age distributions of recovered (i.e., immune against reinfection) individuals were used for the initial population at the start of vaccination. We simulated the model with a 10,000 population for a time horizon of one year to evaluate the impact of DSD vaccination compared with the recommended schedule of 28-day interval for Moderna vaccines and 21-day interval for Pfizer-BioNTech vaccines. For the results presented here, outcomes of infections, hospitalizations, and deaths were averaged over 1000 independent replications of each scenario. Credible intervals (CrI) at the 5% significance level were generated using the bias-corrected and accelerated bootstrap method (with 500 replications).

## Results

### DSD vaccination without waning efficacy of the first dose

When the efficacy of the first dose did not wane for up to 18 weeks after being administered, we found that the DSD strategy with the daily rate of 30 vaccine doses per 10,000 population averted more infections, hospitalizations, and deaths, compared to the recommended schedules for both Moderna and Pfizer-BioNTech vaccines (Figure 2). The largest reduction of severe outcomes was achieved with a 12-15 week delay in administering the second dose. At 20% pre-existing immunity, for example, a 12-week DSD strategy with mean efficacy of Moderna vaccines would avert an additional 0.85 (95% CrI: 0.62 - 1.07) hospitalizations and 0.41 (95% CrI: 0.33 - 0.52) deaths per 10,000 population, compared to the recommended vaccination schedule (Figure 2B-2C). We observed similar benefits of a DSD strategy for Pfizer-BioNTech vaccines, averting 0.74 (95% CrI: 0.48 - 1.04) hospitalizations and 0.41 (95% CrI: 0.31 - 0.54) deaths per 10,000 population in a 12-week delay scenario. As the daily number of vaccine doses increases, the maximum benefits of a DSD strategy in averting hospitalizations and deaths would be achieved with a shorter delay in administering the second dose (Figure 2E-2F).

**Figure 2.**
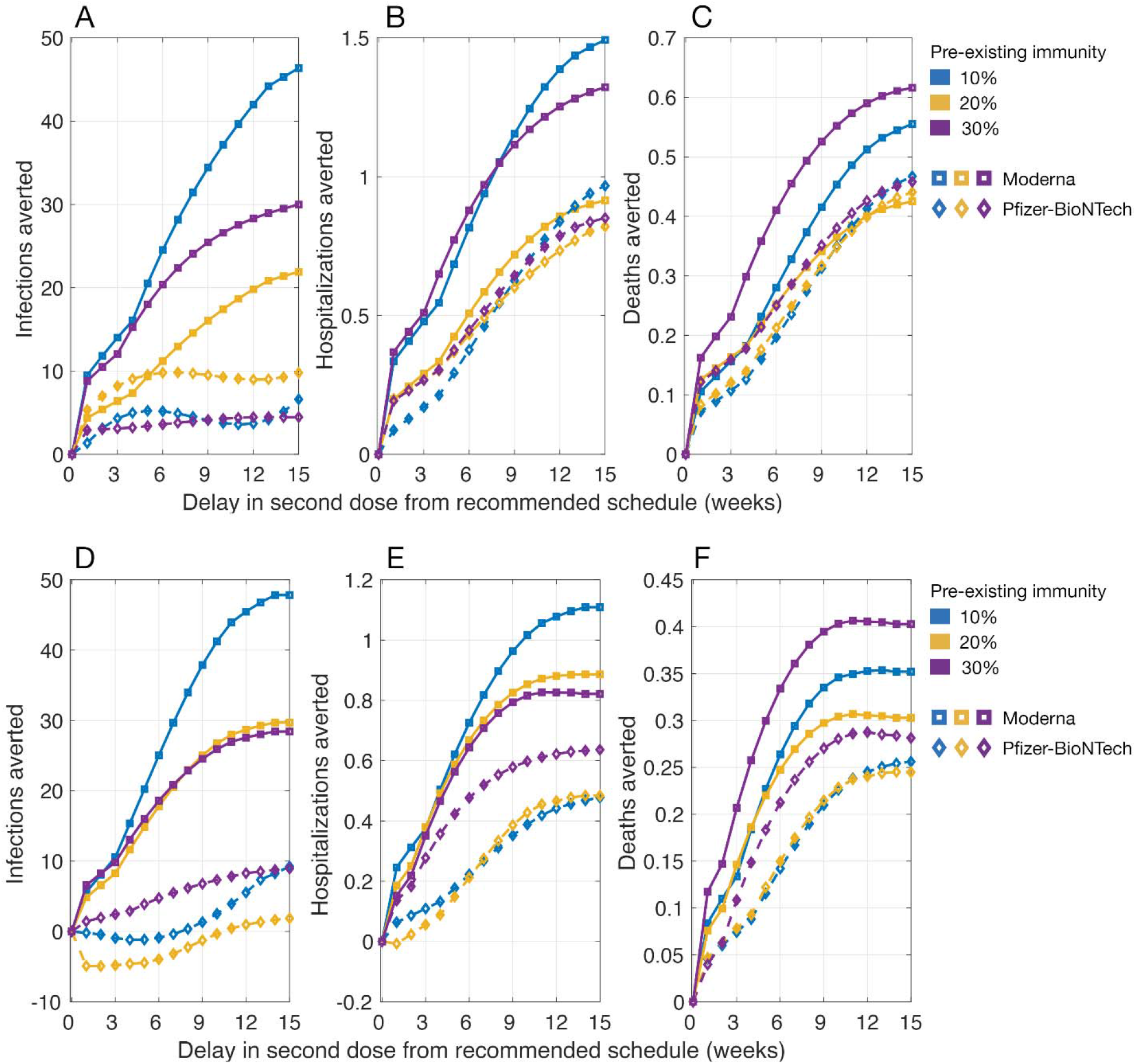
Projected number of infections, hospitalizations, and deaths averted per 10,000 population in a DSD vaccination program compared to the recommended schedule of two-doses of Moderna (with a 28-day interval) and Pfizer-BioNTech (with a 21-day interval) vaccines. The daily vaccination rate was (A,B,C) 30 doses and (D,E,F) 45 doses per 10,000 population. Vaccine efficacy was set to the mean of estimated ranges (Figure 1) without waning of the first-dose efficacy prior to the administration of the second dose.

When simulating the model with upper bounds of vaccine efficacy, we found that the benefits of a DSD strategy in terms of reducing infections, hospitalizations, and deaths were comparable to those obtained in scenarios with mean efficacy for both Moderna and Pfizer-BioNTech vaccines (Appendix, Figure A1). However, when vaccine efficacy was at the lower bounds of estimated ranges, there was no clear advantage with a DSD strategy compared to the recommended schedules in terms of reducing infections (Appendix, Figure A2). While both vaccines averted more hospitalizations and deaths in a DSD strategy, Moderna vaccines outperformed Pfizer-BioNTech vaccines in all scenarios of pre-existing immunity at the lower bounds of vaccine efficacy. The largest reduction of severe outcomes was achieved with a 9-15 week delay in the second dose (Appendix, Figure A2). These benefits are due to the prioritization of elderly and individuals with comorbidities receiving the first dose; thus, increasing their vaccine coverage and reducing severe outcomes among these high-risk individuals.

### DSD vaccination with waning efficacy of the first dose

We found that Moderna vaccines in a DSD strategy averted more infections compared to the recommended schedule of 28 days between doses (Figure 3). However, there was no advantage of DSD using Pfizer-BioNTech vaccines in reducing infections. Both vaccines averted more hospitalizations and deaths with DSD. The largest reduction of hospitalizations and deaths using Moderna vaccines was still achieved with a 12-15 week delay in administering the second dose. With Pfizer-BioNTech vaccines, the highest benefits in reducing the same outcomes would be attained with a shorter delay of 6-12 weeks in a DSD strategy. Overall, Moderna vaccines outperformed Pfizer-BioNTech vaccines with regards to achieving the maximum benefits with a DSD strategy. For example, with 20% pre-existing immunity and a daily vaccination rate of 30 doses, Moderna vaccines averted an additional 0.72 (95% CrI: 0.54 - 0.96) hospitalizations per 10,000 population with a 12-week DSD strategy (Figure 3B). For Pfizer-BioNTech vaccines, this maximum benefit was achieved with a 9-week DSD which averted 0.44 (95% CrI: 0.16 - 0.72) hospitalizations. Similarly, we projected that Moderna and Pfizer-BioNTech vaccines would avert an additional 0.39 (95% CrI: 0.29 - 0.49) and 0.26 (95% CrI: 0.16 - 0.39) deaths per 10,000 population with a 12-week delay of administering the second dose, respectively (Figure 3C).

**Figure 3.**
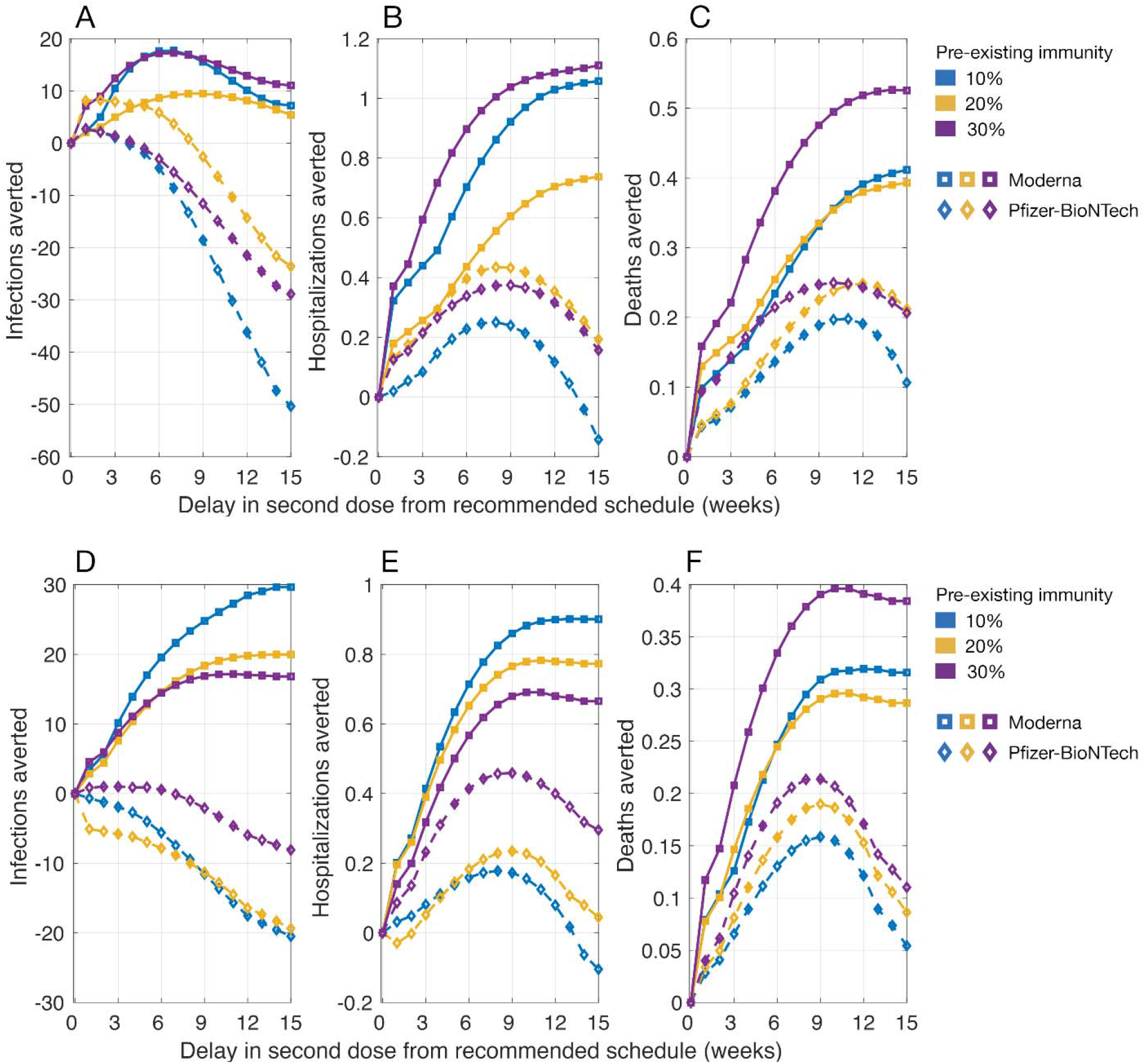
Projected number of infections, hospitalizations, and deaths averted per 10,000 population in a DSD vaccination program compared to the recommended schedule of two-doses of Moderna (with a 28-day interval) and Pfizer-BioNTech (with a 21-day interval) vaccines. The daily vaccination rate was (A,B,C) 30 doses and (D,E,F) 45 doses per 10,000 population. Vaccine efficacy was set to the mean of estimated ranges (Figure 1), and the waning rate of first-dose efficacy was 5% per week, starting from week 7 after the first dose prior to the administration of the second dose.

When the daily vaccination rate increased to 45 doses, we observed similar outcomes of a DSD strategy with Moderna vaccines outperforming Pfizer-BioNTech vaccines in the corresponding scenarios (Figure 3D-3F). The largest benefits of Moderna vaccines in terms of averting hospitalizations and deaths were achieved with a shorter delay of 9-12 weeks in administering the second dose, but still outperformed those obtained using Pfizer-BioNTech vaccines in all scenarios of pre-existing immunity (Figure 3E,3F).

When simulating the model with upper bounds of vaccine efficacy with waning, we found that the performance of a DSD strategy in terms of reducing infections, hospitalizations, and deaths was qualitatively similar to those obtained in scenarios with mean efficacy for both Moderna and Pfizer-BioNTech vaccines (Appendix, Figure A5). However, when vaccine efficacy was at the lower bounds of estimated ranges, the impact of a DSD strategy was reduced significantly in both vaccines (Appendix, Figure A6). There was no advantage in reducing infections with DSD compared to the recommended schedules. We found that, while the performance of a DSD strategy in averting hospitalization and deaths depends on the level of pre-existing immunity, Moderna vaccines still outperformed Pfizer-BioNTech vaccines in most scenarios of pre-existing immunity with a delay of longer than 6 weeks from the recommended schedules (Appendix, Figure A6).

## Discussion

Vaccination can have a substantial impact on mitigating COVID-19 outbreaks [46]. However, vaccine distribution in the US did not meet the initial goal set by federal officials due to significant shortfalls in distribution [47]. Challenges with vaccine supply and rollout, coupled with a deadly wave of outbreaks that overwhelmed hospitals [48–50], and the emergence of highly transmissible SARS-CoV-2 variants [12,51,52], sparked a debate as to whether available vaccines should be used to rapidly increase the coverage with the first dose [14–17], or be distributed according to tested schedules. While the US has committed to delivering the second dose on time for those who receive the first dose [53], a few countries have approved guidelines for DSD, including UK and Canada to defer the second dose by up to 12 and 16 weeks, respectively [54,55].

In this study, we evaluated whether deferral of the second dose beyond the recommended schedules of 3 and 4 weeks for Pfizer-BioNTech and Moderna vaccines, respectively, could improve the effectiveness of vaccination programs in reducing infections, hospitalizations, and deaths. We found that if the efficacy of the first dose did not wane until the administration of the second dose, then the DSD strategy will be more effective than the recommended schedules for both Pfizer-BioNTech and Moderna vaccines, achieving maximum benefits with a delay of 12-15 weeks. If the efficacy of the first dose wanes over time, our results show that delaying the second dose of Moderna vaccines could prevent more infections, hospitalizations, and deaths compared to the recommended 4-week interval between the two doses. The maximum benefits for averting severe outcomes were achieved with a DSD of 9-15 weeks. A DSD strategy with Pfizer-BioNTech vaccines beyond the 3-week tested schedule, on the other hand, may lead to a higher number of infections compared to the recommended schedule, if the first-dose efficacy waned over time. However, depending on the level of pre-existing immunity, additional hospitalizations and deaths could be averted with DSD as a result of vaccine prioritization for individuals at higher risk of severe outcomes. While our study aimed to compare the outcomes of vaccination between the recommended schedules and DSD, we note that the reduction in disease burden by DSD strategy would be even higher when compared to a scenario of no vaccination.

Our results are based on available evidence and estimated vaccine efficacy in published studies of clinical trials, FDA briefing documents, and vaccination campaigns [3,4,6,22–24]. Key data on the durability of vaccine-induced immunity following the first (if the second dose is delayed) and second doses, and the rate of temporal decline of immunity post-vaccination are still lacking. Our model assumptions were conservative and based on limited empirical evidence available thus far. For example, in the base-case scenario, we assumed that the estimated protection efficacy of vaccines against infection and symptomatic/severe disease remained intact until the administration of the second dose. Further, we assumed that the protection efficacies after two doses in a DSD strategy will be the same as those estimated with two doses in schedules tested in clinical trials. As sensitivity analyses, we also considered scenarios in which vaccine efficacy of the first dose waned over time when the second dose was delayed. Currently, there are no data that quantify the decline of vaccine-induced immunity under different schedules of a DSD strategy. However, as clinical investigations on vaccine performance continue and more estimates on population-wide effectiveness of vaccination campaigns become available, our assumptions may need to be revised. Should any future evidence alter these assumptions, further analyses would be warranted and conclusions of our study might change.

Our findings highlight two important parameters in the evaluation of vaccination programs with DSD. First and foremost is the durability of vaccine efficacy [20,21], which requires clinical and epidemiological studies monitoring vaccinated individuals for several weeks after inoculation with the first dose. Second is the ability of vaccines to block transmission. In addition to these parameters, vaccine supply and many other factors such as the potential for the emergence of vaccine-resistant strains under low individual-level protection; public confidence in vaccines; risk behaviour of individuals following vaccination; and the possibility of a drop in uptake of the second dose with a delay significantly longer than the recommended schedules, would be important considerations in public health decision-making regarding DSD vaccination [19]. However, given the relatively high vaccine efficacies estimated after the first dose against severe outcomes (i.e., over 50% in most end-points), broader population-level protection would be expected to further reduce the disease burden, even with limited vaccine supplies in the near term. When racing against a burgeoning outbreak, our results show that prioritizing vaccine coverage with rapid distribution of the first dose would be critical to mitigating adverse outcomes and allow the healthcare system to also address non-COVID-19 medical needs of the population. In the case of low incidence, it would still be important to accelerate vaccination with the first dose to protect the maximum number of individuals ahead of any outbreak surge.

## Data Availability

The computational system and parameters are available under an open source license at: https://github.com/thomasvilches/delayed_dose.

## Reproducibility Statement

The computational system and parameters are available under an open-source license at: https://github.com/thomasvilches/delayed_dose.

