## Supplemental File for "Evaluation of COVID-19 vaccination strategies with a delayed second dose"

This appendix provides further details of model parameterization, and additional results for comparing a DSD strategy with the recommended schedule of vaccination for both Pfizer-BioNTech and Moderna vaccines, corresponding to the efficacy of vaccines against infection.

**Table A1.** Mixing patterns and the daily number of contacts derived from empirical observations. Daily numbers of contacts were sampled from negative binomial distributions for different scenarios.

| Age group | Proportion of contacts between age groups | | | | | No. of daily contacts without self-isolation | No. of daily contacts for self-isolated individuals |
|---|---|---|---|---|---|---|---|
| | 0-4 | 5-19 | 20-49 | 50-65 | 65+ | Mean (SD) | Mean (SD) |
| 0-4 | 0.2287 | 0.1839 | 0.4219 | 0.1116 | 0.0539 | 10.21 (7.65) | 2.86 (2.14) |
| 5-19 | 0.0276 | 0.5964 | 0.2878 | 0.0591 | 0.0291 | 16.793 (11.7201) | 4.70 (3.28) |
| 20-49 | 0.0376 | 0.1454 | 0.6253 | 0.1423 | 0.0494 | 13.795 (10.5045) | 3.86 (2.95) |
| 50-65 | 0.0242 | 0.1094 | 0.4867 | 0.2723 | 0.1074 | 11.2669 (9.5935) | 3.15 (2.66) |
| 65+ | 0.0207 | 0.1083 | 0.4071 | 0.2193 | 0.2446 | 8.0027 (6.9638) | 2.24 (1.95) |

**Table A2.** Description of model parameters and their estimates.

| Description | 0–4 | 5–19 | 20–49 | 50–64 | 65–79 | 80+ | Source |
|---|---|---|---|---|---|---|---|
| Transmission probability per contact during presymptomatic stage | Depending on the level of (herd immunity) 0.042 (10%), 0.0465 (20%), 0.054 (30%) | | | | | | Calibrated to R=1.2 [56] |
| Incubation period (days) | LogNormal(shape: 1.434, scale: 0.661) | | | | | | [31] |
| Asymptomatic period (days) | Gamma(shape: 5, scale: 1) | | | | | | Derived from [33,34] |
| Presymptomatic period (days) | Gamma(shape: 1.058, scale: 2.174) | | | | | | Derived from [29,32] |
| Infectious period from onset of symptoms (days) | Gamma(shape: 2.768, scale: 1.1563) | | | | | | Derived from [33] |
| Proportion of infections that are asymptomatic | 0.30 | 0.38 | 0.33 | 0.33 | 0.19 | 0.19 | [57–59] |
| Proportion of symptomatic cases that exhibit mild symptoms | 0.95 | 0.90 | 0.85 | 0.60 | 0.20 | 0.20 | [25,35] |
| Proportion of cases hospitalized with one or more comorbidities | 37.6% | | | | | | [37,38] |
| Non-ICU | 67% | | | | | | |
| ICU | 33% | | | | | | |
| Proportion of cases hospitalized without any comorbidities | 9% | | | | | | [37,38] |
| Non-ICU | 75% | | | | | | |
| ICU | 25% | | | | | | |
| Length of non-ICU stay (days) | Gamma(shape: 4.5, scale: 2.75) | | | | | | Derived from [39,40] |
| Length of ICU stay (days) | Gamma(shape: 4.5, scale: 2.75) + 2 | | | | | | Derived from [39,40] |

**Table A3.** Vaccination coverage of different age groups over a one-year time horizon.

| Age group | 0-17 | 18-49 | 50-64 | 65-79 | 80+ |
|---|---|---|---|---|---|
| Vaccination coverage | 0% | 59% | 63% | 94% | 96% |

**Results with vaccine efficacy set at upper (Figure A1) and lower (Figure A2) bounds of the estimated ranges without waning of the first-dose efficacy.**

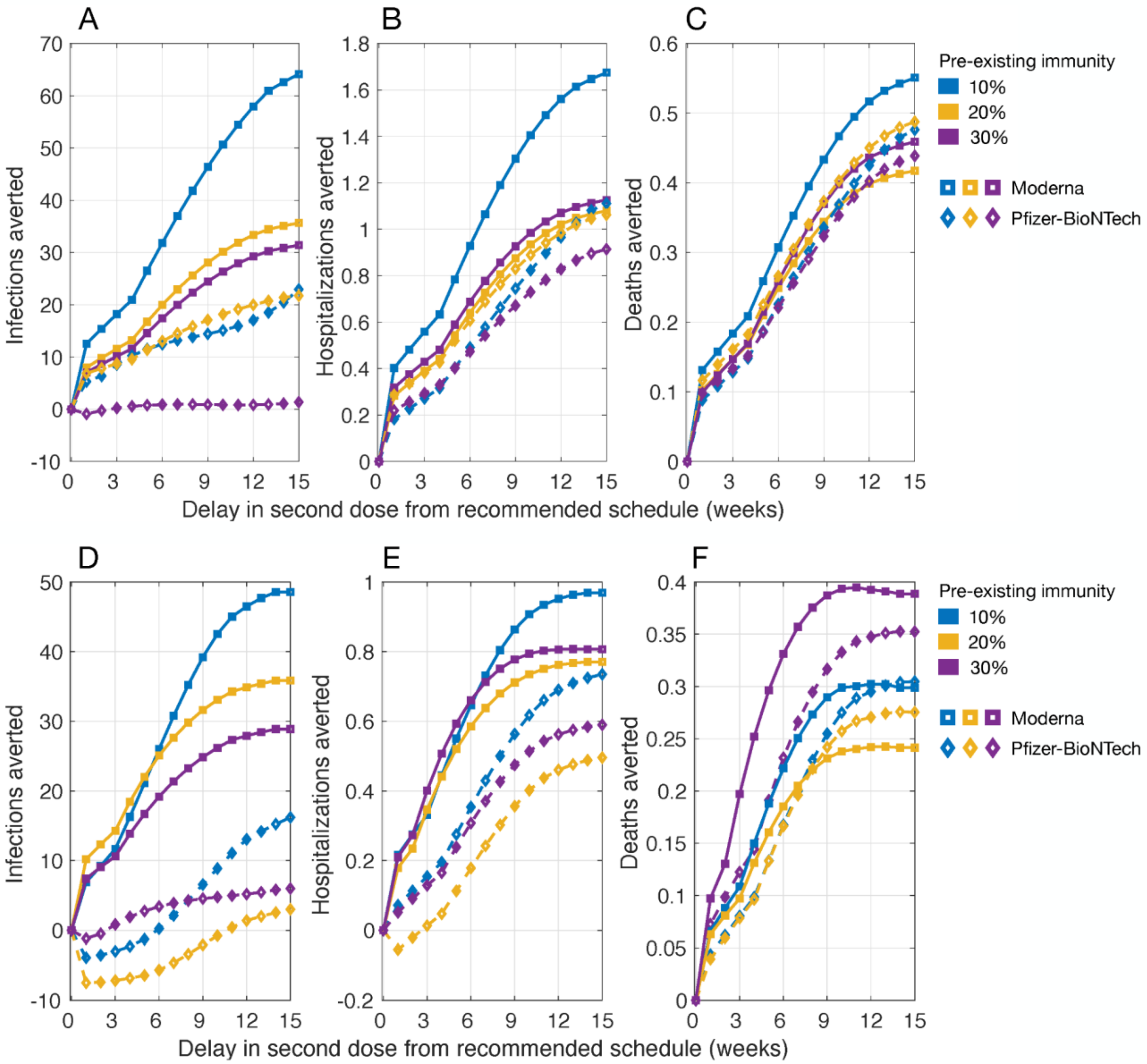


**Figure A1.** Projected reduction of infections, hospitalizations, and deaths for a DSD vaccination program compared to the recommended schedule of two-doses of Pfizer-BioNTech (with a 21-day interval) and Moderna (with a 28-day interval) vaccines. The daily vaccination rate was (A,B,C) 30 doses and (D,E,F) 45 doses per 10,000 population. Vaccine efficacy was set to the upper bound of estimated ranges (Main Text, Figure 1) without waning of first-dose efficacy prior to the administration of the second dose.

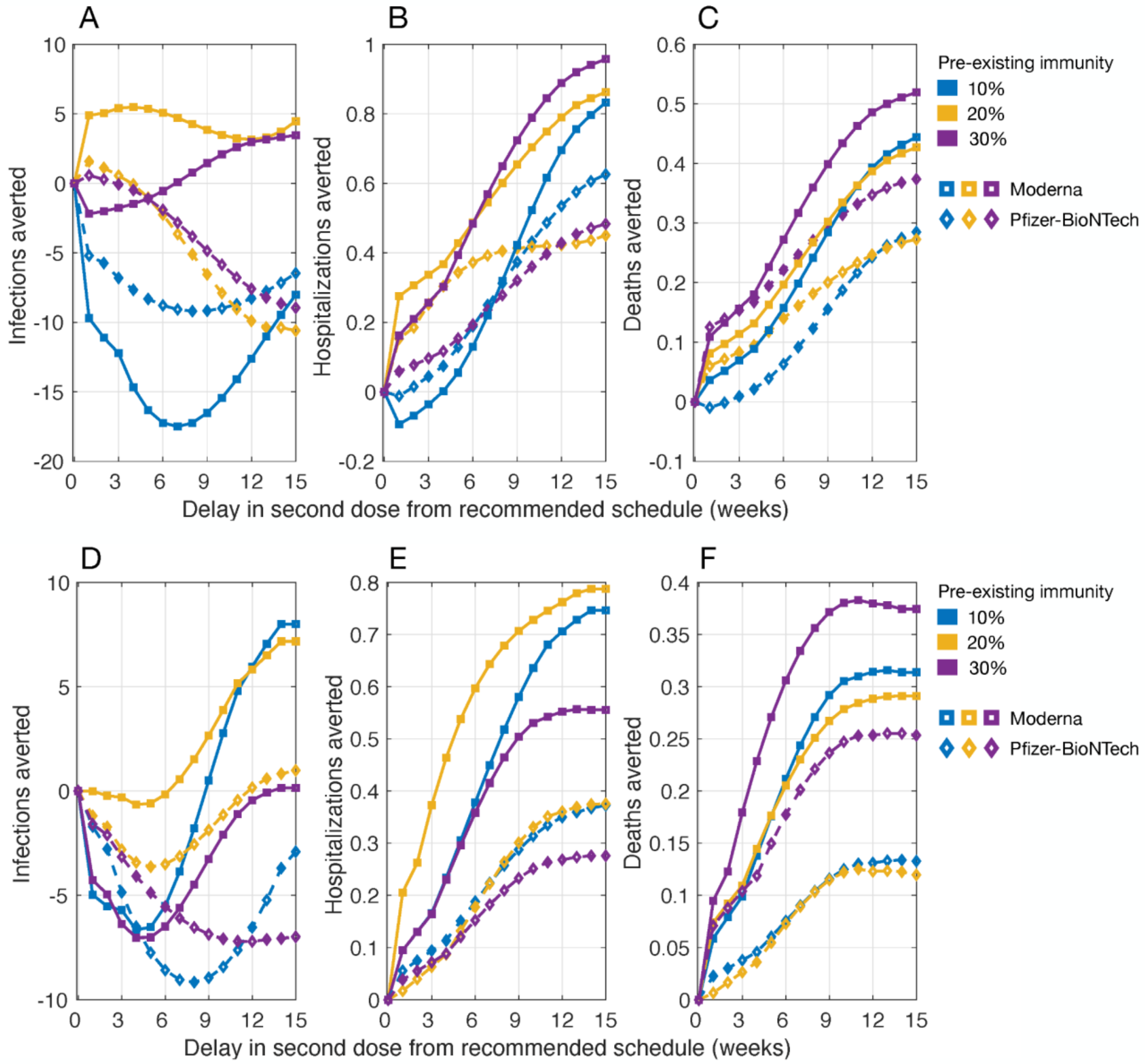


**Figure A2.** Projected reduction of infections, hospitalizations, and deaths for a DSD vaccination program compared to the recommended schedule of two-doses of Pfizer-BioNTech (with a 21-day interval) and Moderna (with a 28-day interval) vaccines. The daily vaccination rate was (A,B,C) 30 doses and (D,E,F) 45 doses per 10,000 population. Vaccine efficacy was set to the lower bound of estimated ranges (Main Text, Figure 1) without waning of first-dose efficacy prior to the administration of the second dose.

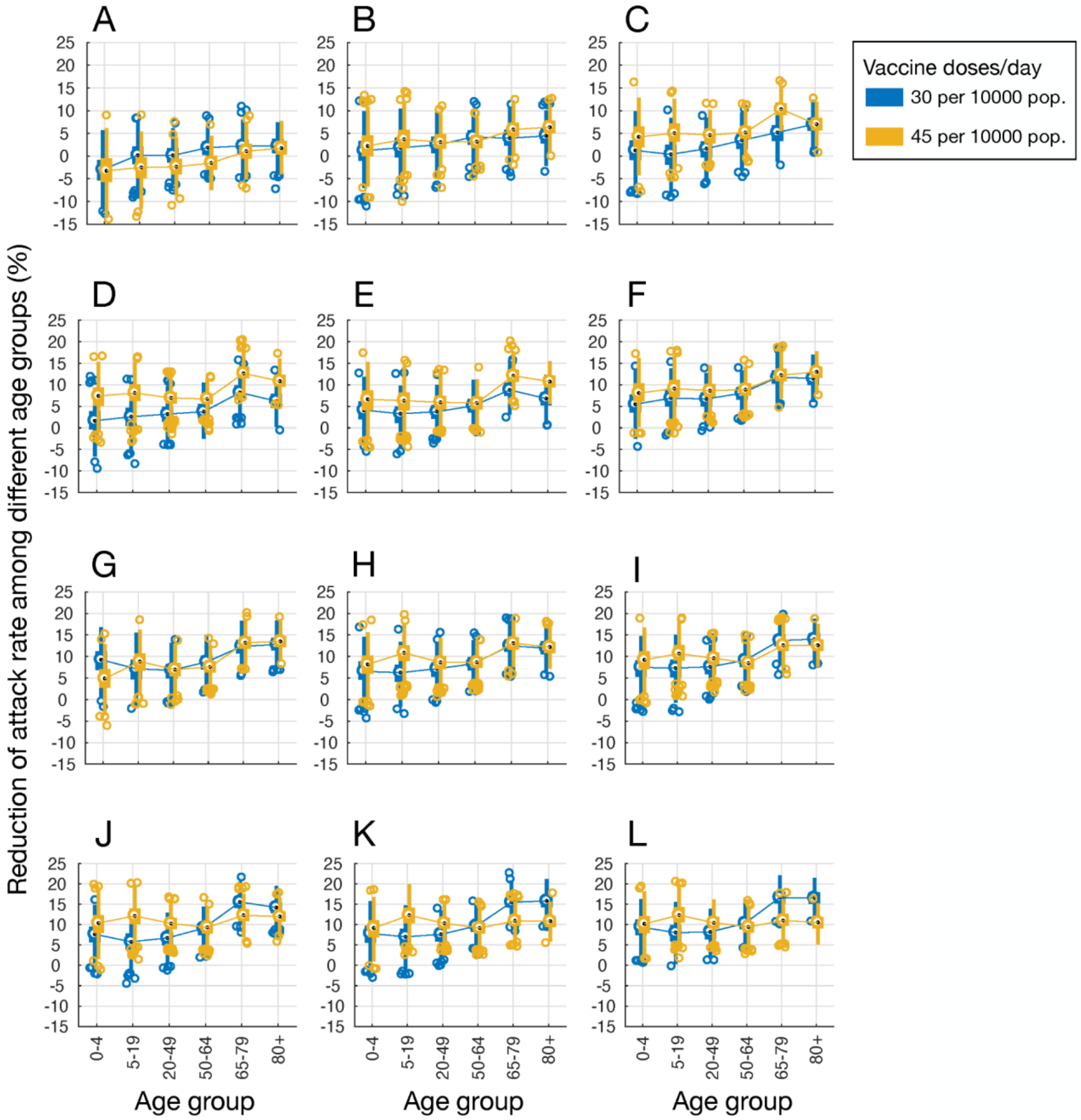


**Figure A3.** Projected reduction of attack rates among different age groups in a DSD strategy with Moderna vaccines. The level of pre-existing immunity was 20% and vaccine efficacy set at the mean values of estimated ranges. Panels A to L correspond to the delay of 1 to 12 weeks in administering the second dose from the recommended schedule, without waning efficacy of the first-dose.

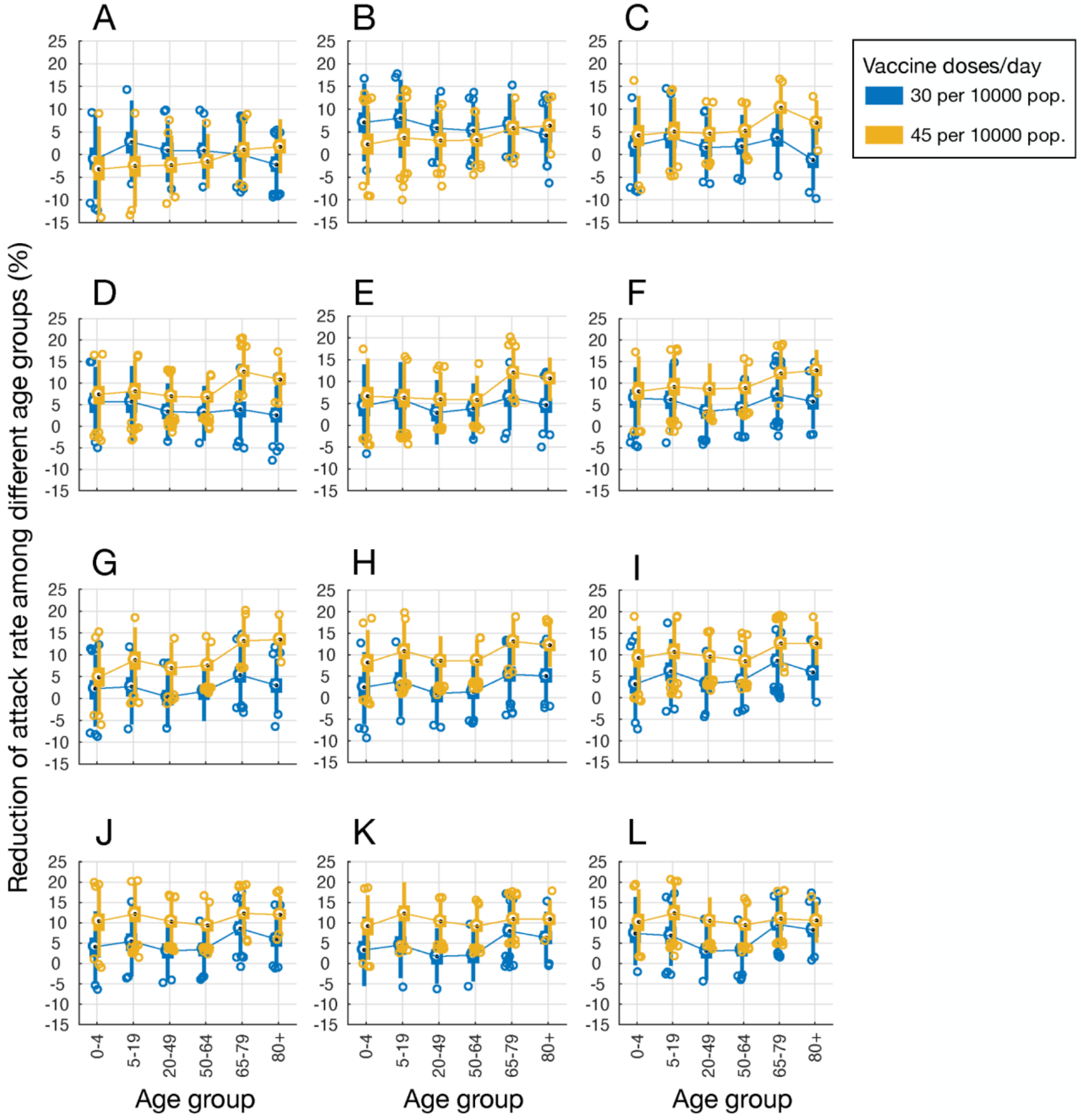


**Figure A4.** Projected reduction of attack rates among different age groups in a DSD strategy with Pfizer-BioNTech vaccines. The level of pre-existing immunity was 20% and vaccine efficacy set at the mean values of estimated ranges. Panels A to L correspond to the delay of 1 to 12 weeks in administering the second dose from the recommended schedule, without waning efficacy of the first-dose.

**Results with vaccine efficacy set at upper (Figure A5) and lower (Figure A6) bounds of the estimated ranges with waning of the first-dose efficacy.**

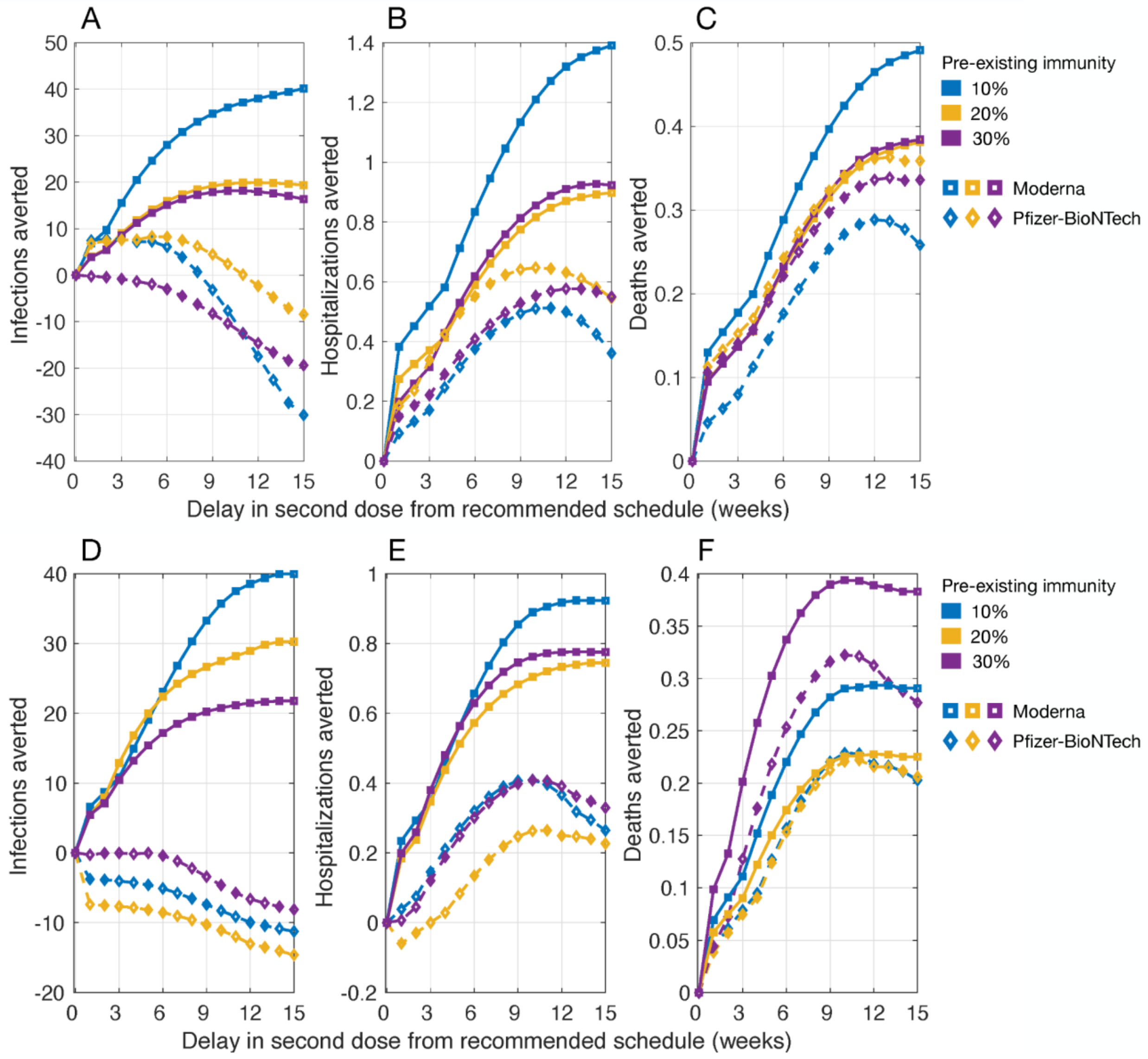


**Figure A5.** Projected reduction of infections, hospitalizations, and deaths for a DSD vaccination program compared to the recommended schedule of two-doses of Pfizer-BioNTech (with a 21-day interval) and Moderna (with a 28-day interval) vaccines. The daily vaccination rate was (A,B,C) 30 doses and (D,E,F) 45 doses per 10,000 population. Vaccine efficacy was set to the upper bound of estimated ranges (Main Text, Figure 1), and the waning rate of first-dose efficacy was 5% per week, starting from week 7 after the first dose prior to the administration of the second dose.

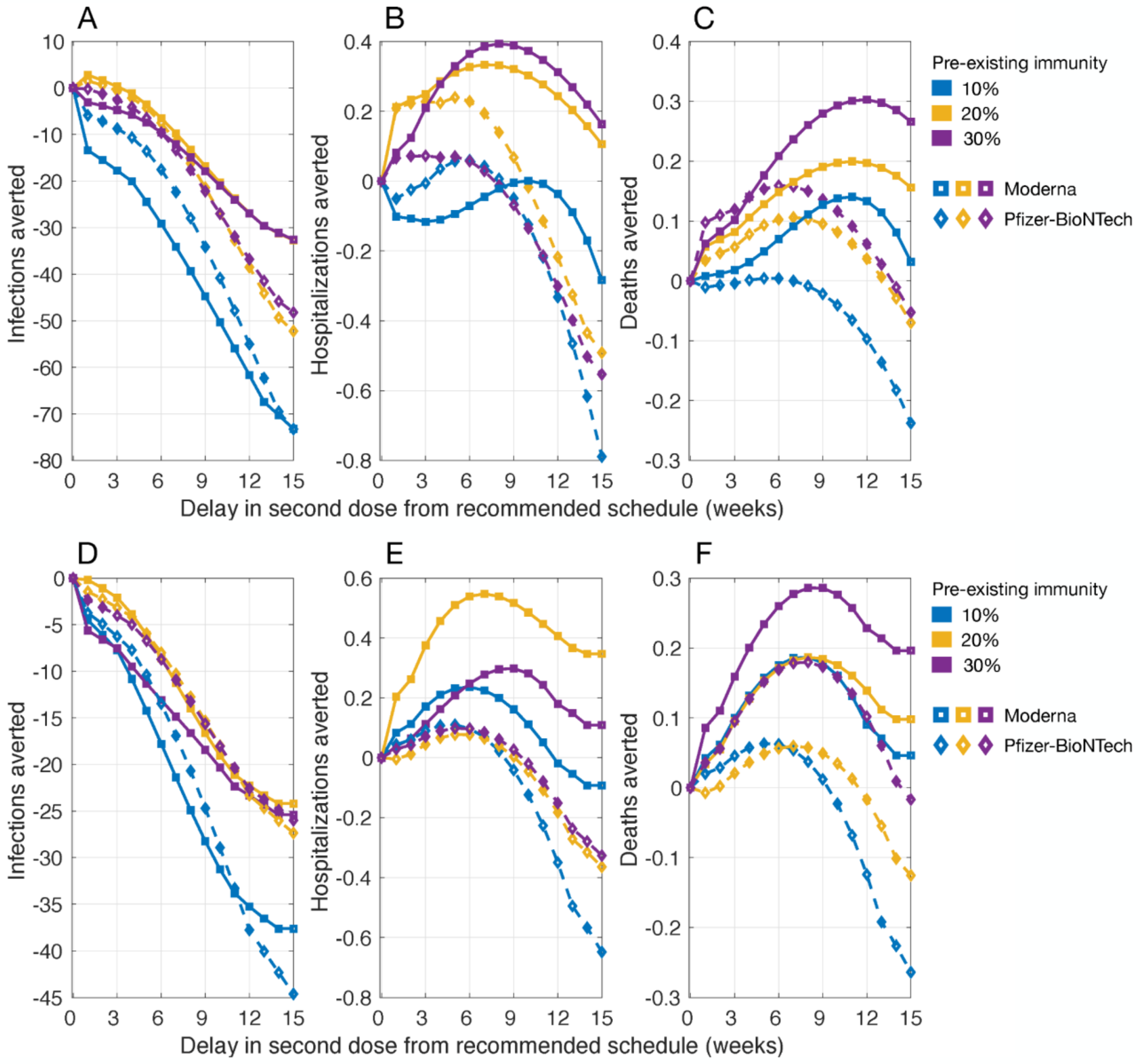


**Figure A6.** Projected reduction of infections, hospitalizations, and deaths for a DSD vaccination program compared to the recommended schedule of two-doses of Pfizer-BioNTech (with a 21-day interval) and Moderna (with a 28-day interval) vaccines. The daily vaccination rate was (A,B,C) 30 doses and (D,E,F) 45 doses per 10,000 population. Vaccine efficacy was set to the lower bound of estimated ranges (Main Text, Figure 1), and the waning rate of first-dose efficacy was 5% per week, starting from week 7 after the first dose prior to the administration of the second dose.
